# Clinical, immunological and genomic characterization of asymptomatic and symptomatic cases with SARS-CoV-2 infection, India

**DOI:** 10.1101/2021.05.21.21257211

**Authors:** Sanchari Chatterjee, Ankita Datey, Soumya Sengupta, Arup Ghosh, Atimukta Jha, Safal Walia, Sharad Singh, Sandhya Suranjika, Gargee Bhattacharya, Eshna Laha, Supriya Suman Keshry, Amrita Ray, Sweta Smita Pani, Amol Suryawanshi, Rupesh Dash, Shantibhusan Senapati, Tushar K. Beuria, Gulam Hussain Syed, Punit Prasad, Sunil Raghav, Satish Devadas, Rajeeb Swain, Soma Chattopadhyay, Ajay Parida

**Affiliations:** Institute of Life Sciences Bhubaneswar, India; Regional Center for Biotechnology, Haryana, India

## Abstract

**Background:** The current global pandemic of Coronavirus disease 2019 (COVID-19), caused by SARS-CoV-2 led to the investigation with clinical, biochemical, immunological and genomic characterization from the patients to understand the pathophysiology of viral infection.

**Methods:** Samples were collected from six asymptomatic and six symptomatic SARS-CoV-2 confirmed hospitalized patients in Bhubaneswar, Odisha, India. Clinical details, biochemical parameters, treatment regime were collected from hospital, viral load was determined by RT-PCR, levels of cytokines and circulating antibodies in plasma were assessed by Bioplex and isotyping respectively. In addition, the whole genome sequencing of viral strains and mutational analysis were carried out.

**Findings:** Analysis of the biochemical parameters highlighted the increased levels of C-Reactive protein (CRP), lactate dehydrogenase (LDH), serum SGPT, serum SGOT and ferritin in symptomatic patients indicating that patients with higher levels of few biochemical parameters might experience severe pathophysiological complications after SARS-CoV-2 infection. This was also observed that symptomatic patients were mostly with one or more comorbidities, especially diabetes (66.6%). Surprisingly the virological estimation revealed that there was no significant difference in viral load of oropharyngeal (OP) samples between the two groups. This suggests that the viral load in OP sample does not correlate with the disease severity and both asymptomatic and symptomatic patients are equally capable of transmitting the virus. Whereas, viral load was higher in plasma and serum samples of symptomatic patients suggesting that the development of clinical complications is mostly associated to high viral load in plasma and serum. This also demonstrated that the patients with high viral load in plasma and serum samples were found to develop sufficient amounts of antibodies (IgG, IgM and IgA). Interestingly, the levels of 7 cytokines (IL-6, IL-.1α, IP-10, IL-8, IL-10, IFN-α2, IL-15) were found to be highly elevated in symptomatic patients, while three cytokines (soluble CD40L, GRO and MDC) were remarkably higher in asymptomatic patients. Therefore, this data suggest that cytokines and chemokines may serve as “predictive indicator” of SARS-CoV-2 infection and contribute to understand the pathogenesis of COVID-19. The whole genome sequence analysis revealed that the current isolates were clustered with 19B, 20A and 20B clades, however acquired 11 additional changes in Orf1ab, spike, Orf3a, Orf8 and nucleocapsid proteins. The data also confirmed that the D614G mutation in spike protein is mostly linked with higher virus replication efficiency and severe SARS-CoV-2 infection as three patients had higher viral load and among them two patients with this mutation passed away.

**Interpretation:** This is the first comprehensive study of SARS CoV-2 patients from India. This will contribute to a better understanding of the pathophysiology of SARS-CoV-2 infection and advance in the implementation of effective disease control strategies.

**Funding:** This study was supported by the core funding of Institute of Life Sciences, Bhubaneswar, Dept of Biotechnology, India.

**Research in context:** *Evidence before this study:* Asymptomatic patients are a source of concern as measures taken to control the spread of the virus are severely impacted by their undetectability. Presently, there is an inadequate information about the characteristics of the asymptomatic and symptomatic patients. The association between SARS-CoV-2 viral load, cytokines and risk of disease progression remains unclear in COVID-19 in Indian scenario. PubMed was searched for articles published up to May, 2021, using the keywords “SARS CoV-2 patients in India”, or “2019 novel coronavirus patients in India”. No published work about the patient’s data on SARS CoV-2 in Indian scenario could be identified.

*Added value of this study:* This investigation highlights the ability of both asymptomatic and symptomatic patients to transmit the virus equally. This study also demonstrates that the D614G mutation in the spike protein is associated with severe SARS-CoV-2 infection and enhance levels of inflammatory markers such as CRP and ferritin which can be predictive biomarkers for critical condition of patients. This is the first comprehensive study of SARS CoV-2 patients from India and will contribute to a better understanding of the pathophysiology of SARS-CoV-2 infection by advancing the implementation of effective disease control strategies.

*Implications of all the available evidence:* The current global pandemic of Coronavirus disease 2019 (COVID-19), caused by SARS-CoV-2 led to the investigation with clinical, biochemical, immunological and viral genome sequencing to understand the pathophysiology of this virus infection. Samples were collected from six asymptomatic and six symptomatic SARS-CoV-2 confirmed hospitalized patients in Bhubaneswar, Odisha, India. This investigation highlights the ability of both asymptomatic and symptomatic patients to transmit the virus equally. This also demonstrated that the D614G mutation is mostly associated with higher virus replication capacity and severe SARS-CoV-2 infection and enhanced levels of inflammatory markers such as CRP and ferritin which are associated with critical conditions of patients. This is the first comprehensive study of SARS CoV-2 patients from India and will contribute to a better understanding of the pathophysiology of SARS-CoV-2 infection by advancing the implementation of competent disease control strategies.

## Introduction

COVID-19 which was initially reported to cause pneumonia and flu-like illness in the city of Wuhan, China has now become considerably more perilous, with global efforts undergoing to combat the deadly disease (1). The beta coronavirus has a spherical enveloped single-stranded positive sense RNA genome with 5´ cap and 3´ poly-A-tail (2).

According to the WHO, 223 countries till date have been affected by the second wave of COVID-19, with 159,319,384 confirmed cases and an increase in number of deaths around the world. Although the vaccination program in India is at its height, total number of vaccinations given are 1,77,214,256. The second wave of COVID-19 cases surpassed the 23,340,938 mark and death toll continues to rise beyond 258,317deaths. As of 13th May 2021, Odisha has 100,313 active cases with 2304 deaths and 64.5 lakhs vaccinated (Ministry of Health and Human Welfare, GOI).

Previous investigations of the SARS-CoV-1 occurrence demonstrated correlation of disease deterioration and viral load in nasopharynx (3,4). Studies have now differentiated SARS-CoV-1 and 2 in terms of clinical manifestations (5), virus shedding (6) and epidemiological aspects (7). However, investigations are required to understand the pathophysiology of SARS-CoV-2 thereby elucidating the association of viremia in different body fluids and disease severity.

Hence, the current investigation was carried out to delineate and compare the clinical manifestations, laboratory findings on viral load, immunological responses, genome sequences, treatment protocol and its outcome of asymptomatic and symptomatic patients infected with SARS CoV-2 to understand the pathophysiology of this virus.

## Materials and Methods

### Study design

In this study, all the patients were admitted to a COVID hospital in Bhubaneswar, Odisha, India in July 2020 and the biological samples including OP swab and, blood were collected. The designed study was approved by Institutional ethics committee and the signed consent forms were taken from the concerned patients. The OP swab sample from the patient was tested immediately to detect the presence of SARS-CoV-2 by RT-PCR. Next, OP swabs were collected from the patients in first (1-3 days), second (5-7 days) and third (8-10 days) phases while blood samples were collected in one time point (1-5 days).

### Data collection

Patient’s demographic and clinical details were obtained from hospital including travel history, treatment details, comorbidities and symptoms. The data for a few patients were missing due to the absence of tests or delayed results.

### Viral RNA extraction

300μl of OP swab/plasma/serum were taken for viral RNA extraction using TAN Bead Maelstrom 4800 as per manufacturer’s instructions. The extracted viral RNA was stored at-80°C until further use.

### qRT-PCR

The qRT-PCR was performed using 5μl of the extracted RNA from samples using the TRUPCR SARS-CoV-2 RT qPCR Kit V-2.0. The human RNase P served as an internal control whereas envelope (E) and nucleocapsid (N) genes were targeted for SARS-CoV-2 amplification.

### Viral copy number determination

The viral copy number was determined as described previously (8). In brief, the standard curve was generated using SARS-CoV-2 N gene specific primers (FP: GTAACACAAGCTTTCGGCAG and RP:GTGTGACTTCCATGCCAATG).

### ELISA

Plasma samples were used to detect the presence of total antibodies (IgM+IgG+IgA) against SARS-CoV-2 by the COVID 19 (IgM+IgG+IgA) microlisa kit (J. Mitra & Co. Pvt. Ltd.) as per manufacturer’s protocol.

### Bioplex

25μl of plasma sample was used to analyse 38 cytokines and chemokines responses using the Bioplex Human Cytokine/Chemokine Magnetic Bead Panel96 Well Plate Assay (EMD Millipore) according to the manufacturer’s instructions.

### Isotyping

To analyse the isotype composition of antibodies in circulation, the plasma of patients was analysed by the ProcartaPlex Human Antibody Isotyping Panels (Invitrogen, Vienna) according to the manufacturer’s instructions.

### Library preparation and whole genome sequencing

The viral amplicon libraries were prepared for whole genome sequencing using the QIAseq FX DNA Library Kit and QIAseq SARS-CoV-2 Primer Panel as per manufacturer’s instructions. The samples were pooled and subjected to IIlumina NextSeq 550 platform in 150x 2 layout, as described before (9).

### Phylogenetic and mutational analysis

The mutational and phylogenetic analysis was carried out using the Nextstrain tool as described before (9). Sequences were aligned along with the WH01 reference strain using the Augur wrapper of MAFFT method to carry out mutational analysis and phylogenetic tree was constructed using the IQTREE2 tool with1,000 bootstrap value (9).

## Statistical analysis

Statistical analysis was performed using the GraphPad Prism software, version 8.0.1. Data were presented as mean ± standard deviation (SD). The non-parametric Mann Whitney U test was used to compare the levels of viral load, cytokines, and antibodies.

## Results

The clinical and biological features of twelve hospitalized patients (asymptomatic and symptomatic) are presented in table 1 and 2. The viral load was assessed after their admission to the COVID hospital (table 3). All the asymptomatic patients (1-6) were recovered and discharged from hospital after 10-11 days. Two symptomatic patients (no. 10 and 11) succumbed to COVID-19 during this period, whereas patient no 9 passed away after getting discharged from hospital. The asymptomatic patients (no.1-6) were identified as COVID-19 positive during contact tracing.

**Table 1.** Demographic details and laboratory findings of SARS CoV-2 infected hospitalized asymptomatic patients.

| Characteristics | Patient 1 | Patient 2 | Patient 3 | Patient 4 | Patient 5 | Patient 6 |
| --- | --- | --- | --- | --- | --- | --- |
| Age range in years | 46-50 | 66-70 | 26-30 | 51-55 | 36-40 | 26-30 |
| Sex | Male | Male | Male | Male | Male | Male |
| Symptoms | Asy | Asy | Asy | Asy | Asy | Asy |
| Comorbidities | None | NA | None | NA | None | None |
| Travel History | Northern part of India | NA | Southern part of India | None | Southern part of India | Southern part of India |
| Medication | Pantoprazole, Azithromycin, Ascorbic acid and Ivermectin | Pantoprazole, Azithromycin, Ascorbic acid and Ivermectin | Pantoprazole, Azithromycin, Ascorbic acid and Ivermectin | Pantoprazole, Azithromycin, Ascorbic acid and Ivermectin | Pantoprazole, Azithromycin, Ascorbic acid and Ivermectin | Pantoprazole, Azithromycin, Ascorbic acid and Ivermectin |
| Duration of stay in Hospital | 11 days | 11 days | 10 days | 11 days | 10 days | 11 days |
| WBC count, 10<sup>3</sup> cells per μL | 3.9 | 5.9 | 5.8 | 05 | 8.7 | 5.8 |
| Neutrophils, differential count(%) | 43 | 43 | 50 | 66 | 71 | 55 |
| Lymphocytes, differential count(%) | 46 | 45 | 38 | 15 | 17 | 36 |
| Monocytes, differential count(%) | 08 | 09 | 09 | 13 | 09 | 07 |
| Eosinophils, differential count(%) | 03 | 03 | 03 | 06 | 03 | 02 |
| Basophils, differential count(%) | 0 | 0 | 0 | 0 | 0 | 0 |
| RBC count, 10<sup>6</sup> per μL | 3.7 | 5.95 | 4.96 | 5.46 | 5.43 | 5.76 |
| **Haemoglobin, gm/dL** | 10.4 | 13.6 | 15 | 14.5 | 14 | 17.4 |
| **MCV, fL/$\mu\text{m}^3$** | 88.1 | 71.4 | 88.7 | 80.2 | 78.8 | 88.4 |
| **Platelet count, $10^3$ per $\mu\text{L}$** | 155 | 250 | 211 | 165 | 93 | 164 |
| **Ferritin, ng/mL** | 115 | 492 | 361 | 115 | 57 | 218 |
| **Serum Bilirubin total, mg per dL** | 0.12 | 0.45 | 1.2 | 0.42 | 0.5 | 0.92 |
| **Serum SGOT, U/L** | 30 | 59 | 38 | 38 | 26 | 45 |
| **Serum SGPT, U/L** | 23 | 142 | 39 | 48 | 19 | 42 |
| **Serum alkaline phosphatase, U/L** | 48 | 91 | 89 | 64 | 94 | 127 |
| **Serum Albumin, g/L** | 4.0 | 4.6 | 4.6 | 3.9 | 4.1 | 5 |
| **Serum Globulin, g/L** | 2.7 | 2.7 | 2.5 | 2.3 | 2.4 | 2.7 |
| **Status of the patients** | Recovered | Recovered | Recovered | Recovered | Recovered | Recovered |

Asy- Asymptomatic
NA- Not available

**Table 2.** Demographic details and laboratory findings of SARS CoV-2 infected hospitalized symptomatic patients.

| **Characteristics** | **Patient 7** | **Patient 8** | **Patient 9** | **Patient10** | **Patient 11** | **Patient 12** |
| --- | --- | --- | --- | --- | --- | --- |
| **Age range in years** | 56-60 | 51-55 | 61-65 | 56-60 | 31-35 | 41-45 |
| **Sex** | Female | Male | Male | Male | Male | Female |
| **Symptoms** | Fever,<br>Tiredness,<br>headache and<br>Joint Pain | Dyspnea,<br>myalgia | Dyspnea,<br>Fever,<br>Tiredness,<br>myalgia | Dyspnea,<br>myalgia | Dyspnea,<br>myalgia | Dyspnea,<br>myalgia |
| **Comorbidities** | None | Diabetic | Diabetic | NA | Diabetic | Diabetic |
| **Travel History** | Eastern part of<br>India | NA | None | NA | NA | NA |
| **Medication** | Pantoprazole,<br>Azithromycin,<br>Ascorbic acid<br>and Ivermectin | Pantoprazole,<br>Azithromycin,<br>Ascorbic acid<br>and Ivermectin | Pantoprazole,<br>Azithromycin,<br>Ascorbic acid<br>and Ivermectin | Pantoprazole,<br>Azithromycin,<br>Ascorbic acid<br>and Ivermectin | Pantoprazole,<br>Azithromycin,<br>Ascorbic acid<br>and Ivermectin | Pantoprazole,<br>Azithromycin,<br>Ascorbic acid<br>and Ivermectin |
| **Duration of stay in**<br>**Hospital** | 11 days | 11 days | 11 days | 6 days | 6days | 11 days |
| **WBC count, 10<sup>3</sup>**<br>**cells per μL** | 9.5 | 5.6 | 11 | 3.5 | 6.3 | 5.3 |
| **Neutrophils,**<br>**differential**<br>**count(%)** | 43 | 83 | 91 | 55 | 77 | 66 |
| **Lymphocytes,**<br>**differential**<br>**count(%)** | 36 | 11 | 06 | 30 | 18 | 15 |
| **Monocytes,**<br>**differential**<br>**count(%)** | 10 | 05 | 03 | 08 | 05 | 13 |
| **Eosinophils,**<br>**differential**<br>**count(%)** | 11 | 01 | 00 | 07 | 00 | 06 |
| **Basophils,**<br>**differential**<br>**count(%)** | 00 | 00 | 00 | 00 | 00 | 00 |
| **RBC count, 10<sup>6</sup>**<br>**per μL** | 3.8 | 4.6 | 4.3 | 2.5 | 6.0 | 5.4 |
| **Haemoglobin, gm/dL** | 11.5 | 13 | 12.7 | 8.6 | 13.7 | 14.5 |
| **MCV, fL/$\mu\text{m}^3$** | 94.5 | 83.3 | 84.5 | 93.4 | 69.7 | 80.2 |
| **Platelet count, $10^3$ per $\mu\text{L}$** | 184 | 125 | 218 | 49 | 230 | 165 |
| **CRP, mg/L** | 95 | 60.12 | 52.38 | 18.93 | NA | 89 |
| **D Dimer $\mu\text{g/mL}$** | NA | 2.38 | 0.63 | NA | NA | NA |
| **Ferritin ng/mL** | 105 | 1000 | 492 | 968 | NA | 115 |
| **Serum LDH U/L** | NA | 456.79 | NA | 724.33 | NA | 723 |
| **Serum Bilirubin total, mg/dL** | 0.55 | 0.91 | 0.61 | 5.85 | 0.83 | 0.8 |
| **Serum SGOT, U/L** | 38 | 67 | 56 | 226 | 107 | 49 |
| **Serum SGPT, U/L** | 20 | 120 | 50 | 165 | 128 | 16 |
| **Serum alkaline phosphatase, U/L** | 89 | 134 | 34 | 61 | 85 | 100 |
| **Serum Albumin, gm/dL** | 4.0 | 3.5 | 3.3 | 2.2 | 4.5 | 4.0 |
| **Serum Globulin, gm/dL** | 3.1 | 2.7 | 2.7 | 3.6 | 3.2 | 3.1 |
| **Status of the patients** | Recovered | Recovered | Demised | Demised | Demised | Recovered |

NA- Not available

**Table 3.** Confirmation of COVID-19 by RT-PCR and virus isolation.

| Patient no. | Month of admission | Nature of specimen | E gene | N gene | RNaseP | Virus adaptation |
| --- | --- | --- | --- | --- | --- | --- |
| 1 | July, 2020 | Oropharyngeal swab | 17.5 | 18.6 | 32.2 | No |
| 2 | July, 2020 | Oropharyngeal swab | 21.8 | 23.2 | 26.1 | No |
| 3 | July, 2020 | Oropharyngeal swab | 19.5 | 20.8 | 26.9 | No |
| 4 | July, 2020 | Oropharyngeal swab | 20.5 | 21.9 | 25.4 | No |
| 5 | July, 2020 | Oropharyngeal swab | 19.1 | 20 | 25.2 | No |
| 6 | July, 2020 | Oropharyngeal swab | 21.6 | 23.3 | 25.2 | No |
| 7 | July, 2020 | Oropharyngeal swab | 19.4 | 22.9 | 25 | Yes |
| 8 | July, 2020 | Oropharyngeal swab | 24.1 | 26.2 | 29.2 | No |
| 9 | July, 2020 | Oropharyngeal swab | 16.4 | 18.7 | 25.7 | No |
| 10 | July, 2020 | Oropharyngeal swab | 16.9 | 17.1 | 28.8 | No |
| 11 | July, 2020 | Oropharyngeal swab | 17.5 | 18 | 26.9 | No |
| 12 | July, 2020 | Oropharyngeal swab | 20.3 | 22.5 | 28.4 | No |

The analysis of the biochemical data revealed that all the symptomatic patients had higher level of CRP, 50% of them displayed high amount of serum LDH (∼1.8-2.9 fold) than normal range. Besides, they were detected with higher percentage of serum SGPT (>2 fold), serum SGOT (>2 fold), and almost 83% of the SARS CoV-2 patients were detected with higher level of ferritin than normal range (Table 1 and 2). This data suggests that the patients with higher levels of the above mentioned biochemical parameters might experience severe pathophysiological complications after SARS-CoV-2 infection.

For the treatment of this disease, all the patients were administered with Pantoprazole (before food), Azithromycin (once a day), Ascorbic acid (2 hrs after food) and Ivermectin (2 hrs after food), for a period of 10 days. Asymptomatic patients were discharged from the hospital on day 10. The symptomatic patients were found to have one or more comorbidities, especially diabetes (66.6%) indicating that other physiological conditions (Table 1 and 2) are playing major role in determining the disease pathology.

Post onset of disease, samples were collected within 1-3 days, 5-7 days and 8-10 days. In most of the asymptomatic (except one) and symptomatic patients (except two), the Ct values were increased in OP samples over time. However, there was no significant difference in Ct values of asymptomatic and symptomatic patients (Fig. 1A). This suggests that the viral load in OP sample does not correlate with disease severity. In contrast, the viral load estimation from plasma samples demonstrated that symptomatic patients had high viral load (low Ct value and high viral copy number) as compared to asymptomatic patients (Fig. 1B). The data for viral copy number is not shown. Three symptomatic patients (patient no. 9, 10 and 11) who died of SARS-CoV-2 had significantly higher amount of plasma viremia, compared to those who were discharged from the hospital. All the symptomatic patients (except one) were on ventilator support. The analysis of serum samples demonstrated that the Ct values of serum samples of asymptomatic patients were more [less viral copy number (data not shown)] than the symptomatic patients and this finding correlated well with the Ct values of plasma samples for both asymptomatic and symptomatic patients (Fig. 1C). Hence, the result indicates that development of clinical complications is mostly associated to high viral load in plasma and serum samples.

**Fig. 1.**
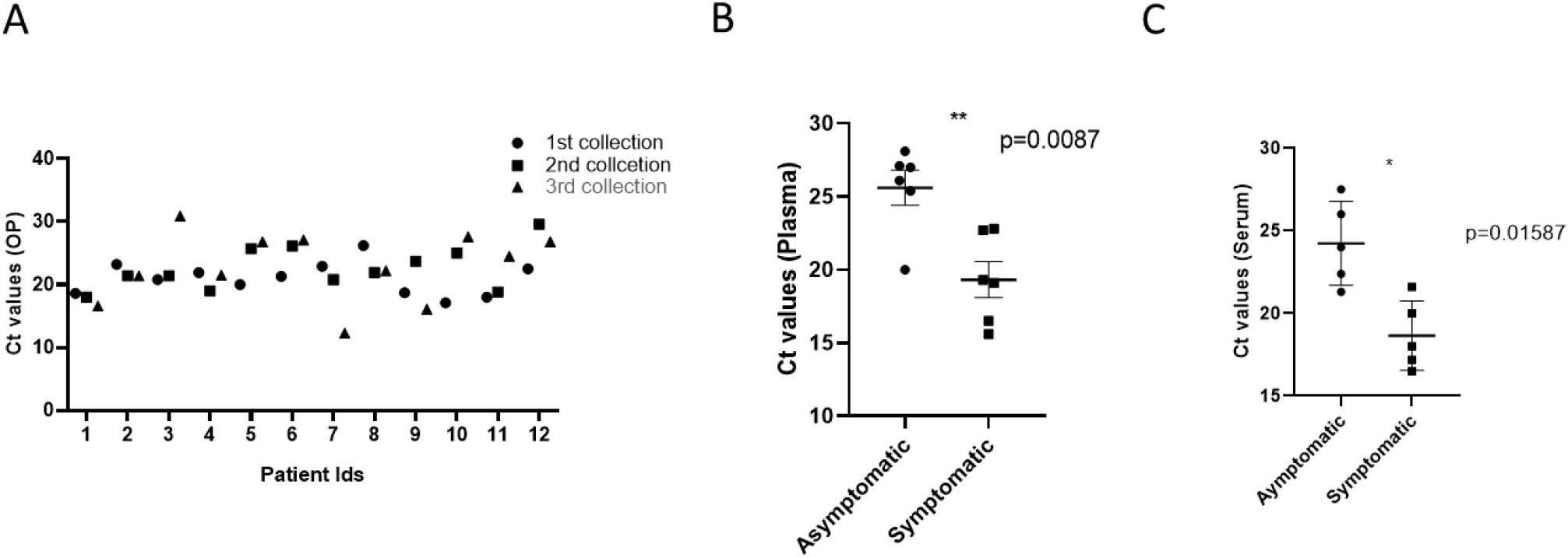
Viral dynamics in patients with asymptomatic and symptomatic disease. (A)ΔCT of OP samples from patients with asymptomatic and symptomatic COVID-19 at different days of disease onset. (B) and (C) ΔCT of plasma and serum samples from asymptomatic and symptomatic COVID-19 patients. The Mann- Whitney (non parametric, two tailed) test was performed. All error bars were SD.

To understand the antibody responses against SARS-CoV-2, virus specific IgM, IgG and IgA antibodies were estimated using plasma samples. Four out of six asymptomatic patients (patient no 2, 4, 5 and 6) along with all symptomatic patients were found to be ELISA positive (with low Ct values in plasma) (Fig. 2). Samples of three healthy individuals were collected as control and were negative for SARS-CoV-2 specific IgM, IgG and IgA antibodies (data not shown). Thus, the data underline that patients with high viral titer developed sufficient amount of antibodies

**Fig. 2.**
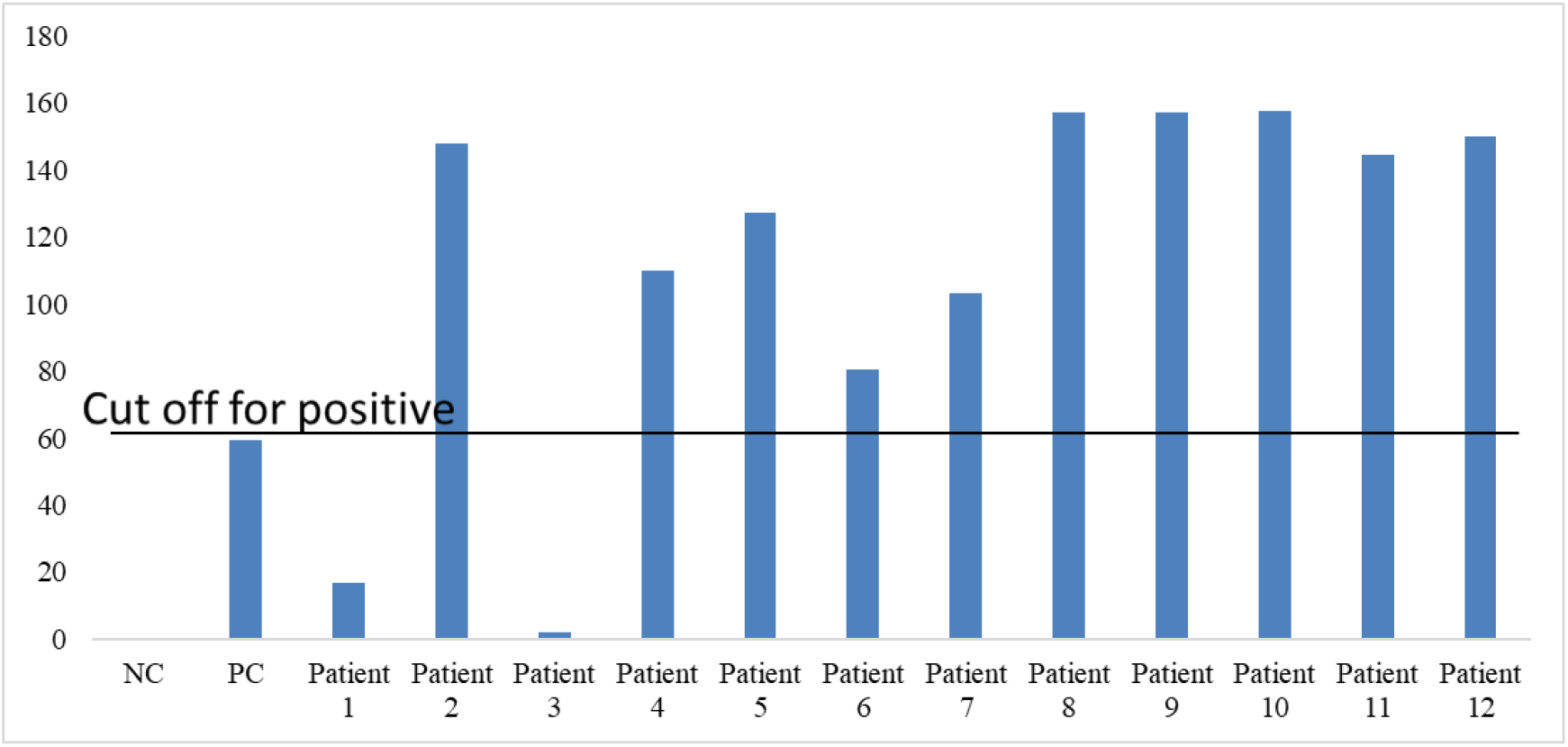
Analysis of SARS CoV-2 specific (IgG+IgM+IgA) antibodies in asymptomatic and symptomatic patients. Bar graph representing antibody units (IgG+IgM+IgA) of COVID-19 patient’s plasma samples, Line depicting the cut off value of positive samples which was 59.3. Negative (NC) and positive (PC) controls delivered by detection kit were included to warrant test validity.

To comprehend infectivity, live virus was isolated from 6 clinical samples. The virus was readily isolated from OP samples from patient 7 (viral load >10^7^ copies/mL) which has highest viral load among all the samples. This indicates the presence of significantly high copy number of virus in this patient.

Next, the differential dynamics of circulating cytokines and chemokines in the SARS-CoV-2 infected asymptomatic and symptomatic patients were investigated. Interestingly, the levels of soluble CD40L, GRO and MDC were higher in asymptomatic patients as compared to the symptomatic. On the other hand, the symptomatic patients showed higher levels of IL-6, IL-1α, IP-10, IL-8, IL-10, IFN-α2 and IL-15 in comparison with the asymptomatic patients (Fig. 3). Whereas, there were no significant difference in the plasma levels of MCP-3, IL-1β, IL-17A, IL-12 p70, eotaxin and TNF. Therefore, this data suggest that cytokines and chemokines may serve as “predictive indicator” of SARS-CoV-2 infection and contribute to understand the pathogenesis of COVID-19.

**Fig. 3.**
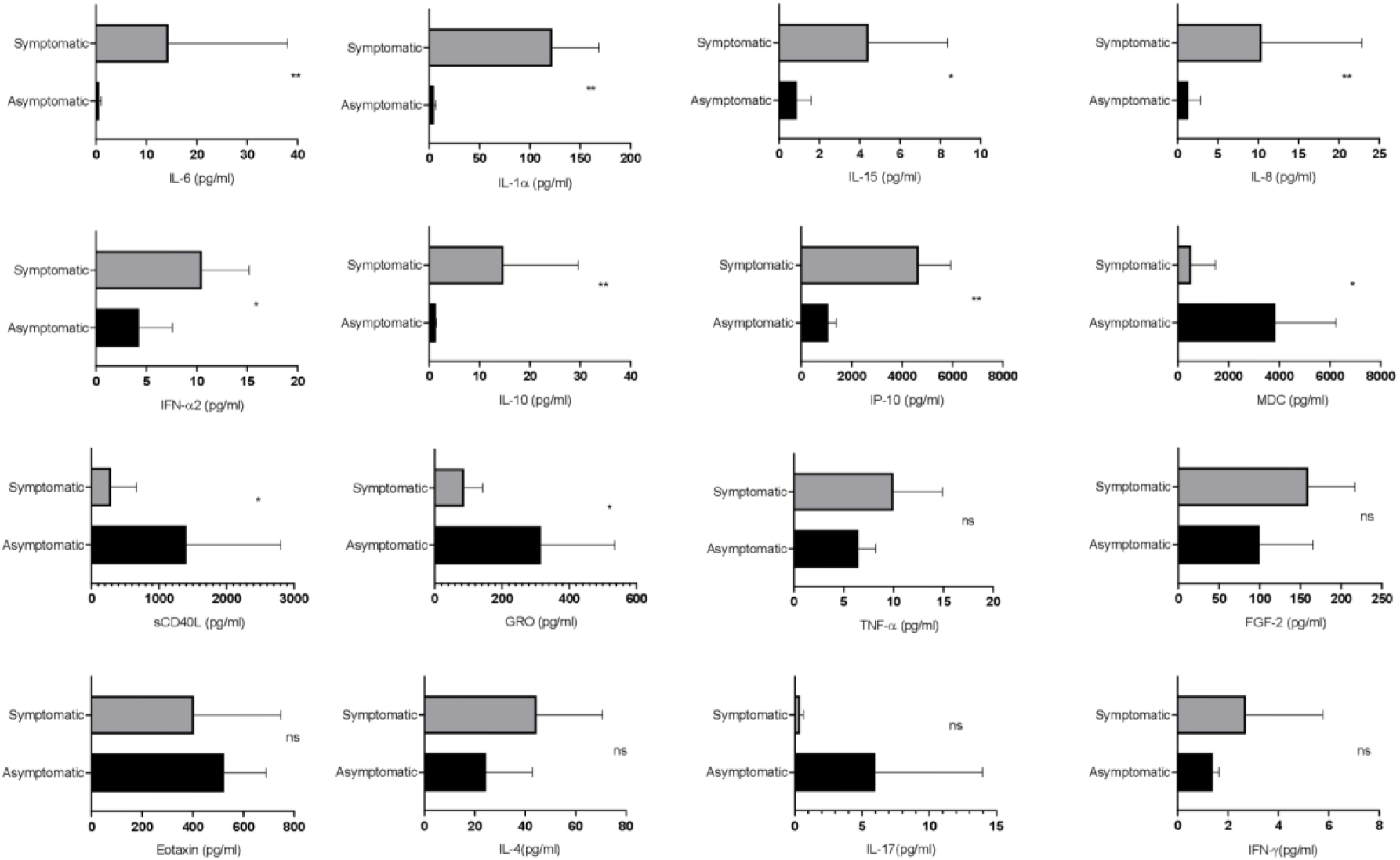
Evaluation of the cytokines responses between asymptomatic and symptomatic COVID-19 patients. The bar diagram depicting the expression levels of cytokines in pg/mL. Cytokines were measured after disease onset between asymptomatic and symptomatic patients. The Mann- Whitney (non parametric, two tailed) test was performed p<0.05 was considered statistically significant (*), and p<0.005 was considered to be very significant (**). ns, not significant. All error bars were SD.

To understand the dynamics of circulating isotypes of antibody in the peripheral blood of SARS-CoV-2 patients, the Bioplex assay was carried out. There was no significant difference between IgA, IgM and IgG4 in the two groups, however, IgE, IgG2, IgG3 and IgG1 were significantly higher in symptomatic than the asymptomatic patients (Fig. 4). This result indicates that inflammatory responses were high in these patients.

**Fig 4.**
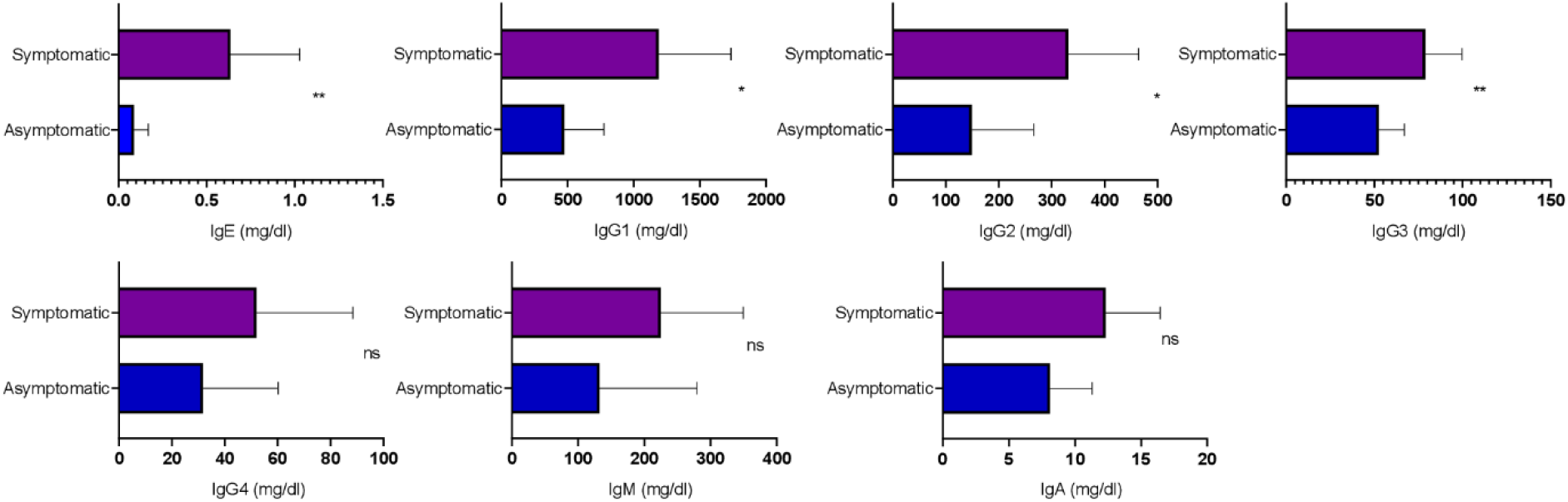
Analysis of the circulating antibodies in asymptomatic and symptomatic COVID-19 patients. Circulating antibody isotypes (IgA, IgM, IgG1, IgG2, IgG3, IgG4 and IgE) were evaluated from COVID -19 positive plasma samples. The Mann- Whitney (non parametric, two tailed) test was performed p<0.05 was considered statistically significant (*), and p<0.005 was considered to be very significant (**). ns, not significant. All error bars were SD.

In the current investigation, whole genome sequencing was also performed for four SARS-CoV-2 strains (from one asymptomatic and three symptomatic patients). All four sequences were deposited to the GISAID database and the accession numbers are listed in Table no.4. The phylogenetic tree was prepared using these isolates and the strains from Wuhan, WH-01 (NC_045512.2), USA, Brazil, Canada, Australia, South Africa, United Kingdom and South Korea were used as references. The phylogenetic analysis revealed presence of three main clades, two (patient no.7 and 9) of the strains clustered to 20A clade whereas the rest two (patient no.1 and 10) strains belonged to 19B and 20B clade (Fig 5).

**Table 4.** Accession IDs of GISAID submissions.

| Patient no. | Virus name | Accession ID | Collection time | Location | Host | Passage | Sequencing technology | Assembly method |
| --- | --- | --- | --- | --- | --- | --- | --- | --- |
| 1 | hCoV-19/India/OR-ILS04/2020 | EPI_ISL_1241861 | July, 2020 | Asia / India / Odisha | Human | Original | Illumina NextSeq 550 | Reference based |
| 7 | hCoV-19/India/OR-ILS06/2020 | EPI_ISL_1242030 | July, 2020 | Asia / India / Odisha | Human | Passage 9 | Illumina NextSeq 550 | Reference based |
| 9 | hCoV-19/India/OR-ILS08/2020 | EPI_ISL_1255389 | July, 2020 | Asia / India / Odisha | Human | Original | Illumina NextSeq 550 | Reference based |
| 10 | hCoV-19/India/OR-ILS09/2020 | EPI_ISL_1255390 | July, 2020 | Asia / India / Odisha | Human | Original | Illumina NextSeq 550 | Reference based |

\* shows the isolates used in this study

**Fig. 5.**
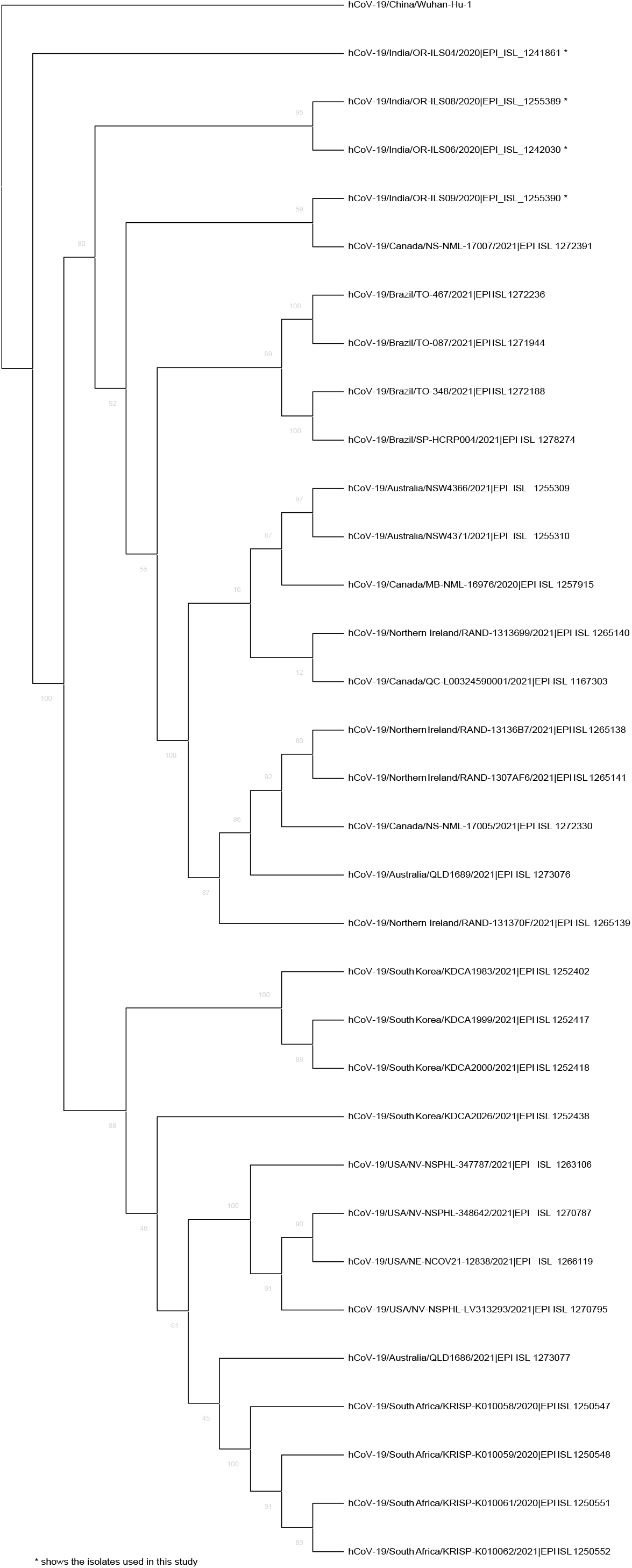
The figure depicts the phylogenetic analysis of the whole genome sequences of four SARS CoV2 strains from human oropharyngeal swab samples. The phylogenetic tree was generated using the Maximum Likelihood method with 1000 bootstrap value by the IQTREE2 tool. The tree was constructed using WH-01 (NC_045512.2) as a reference strain along with four strains from each of the country namely USA, Brazil, Canada, Australia, South Africa, United Kingdom, and South Korea. The sequences for reference strains were retrieved from the GISAID database. The viral isolates are depicted by hCoV-19/country/strain ID/year of isolation/accession number. The bootstrap values are mentioned at major branch points of the tree.

Moreover, the sequence analysis revealed the presence of additional mutations, such as 13 in Orf1ab region, 1 each in spike, Orf3a, Orf8 and nucleocapsid genes. All the patients showed different combinations of mutations (approximately 5-6) that are listed in Table 5.

**Table 5.** Mutational analysis of the four SARS-CoV-2 strains compared to the Wuhan-Hu-1 (NC_045512.2) reference sequence.

| Pos | Gene | Ref | Alt | AA Pos | Ref AA | Alt AA | Patient 1 | Patient 7 | Patient 9 | Patient 10 |
| --- | --- | --- | --- | --- | --- | --- | --- | --- | --- | --- |
| 147 | Orf1ab | C | T | gp01 intergenic_region | - | - | 0 | 0 | 1 | 0 |
| 241 | Orf1ab | C | T | gp01 intergenic_region |  |  | 0 | 0 | 1 | 1 |
| 313 | Orf1ab | C | T | 16 S | Leu | Leu | 0 | 0 | 0 | 1 |
| 2453 | Orf1ab | C | T | 730 M | Leu | Phe | 0 | 0 | 0 | 1 |
| 3037 | Orf1ab | C | T | 924 S | Phe | Phe | 0 | 1 | 0 | 1 |
| 3307 | Orf1ab | G | T | 1014 M | Met | Ile | 0 | 1 | 0 | 0 |
| 5700 | Orf1ab | C | A | 1812 M | Ala | Asp | 0 | 0 | 0 | 1 |
| 8782 | Orf1ab | C | T | 2839 S | Ser | Ser | 1 | 0 | 0 | 0 |
| 10870 | Orf1ab | G | T | 3535 S | Leu | Leu | 0 | 1 | 0 | 0 |
| 11003 | Orf1ab | C | T | 3580 M | His | Tyr | 0 | 1 | 1 | 0 |
| 11230 | Orf1ab | G | T | 3655 M | Met | Ile | 1 | 0 | 0 | 0 |
| 14408 | Orf1ab | C | T | 4715 M | Pro | Leu | 0 | 1 | 1 | 1 |
| 18029 | Orf1ab | C | T | 5922 M | Ale | Val | 0 | 1 | 1 | 0 |
| 23403 | Spike | A | G | 614 M | Asp | Gly | 0 | 1(Low depth)<sup>1</sup> | 1 | 1 |
| 25511 | Orf3a | C | T | 40 M | Ser | Leu | 1 | 0 | 0 | 0 |
| 28167 | Orf 8 | G | A | 92 M | Glu | Lys | 1 | 0 | 0 | 0 |
| 28878 | Nucleocapsid | G | A | 202 M | Ser | Asn | 1 | 0 | 0 | 0 |

<sup>1</sup> Low depth, dp<20

Mutational analysis of amino acid unveiled a total of 11 missense mutations in Orf1ab region whereas 4 synonymous mutations were noticed in spike, nucleocapsid, Orf3a, and Orf8 regions. Interestingly, D614G mutation of spike protein was observed in patient number 7, 9 and 10 and the analysis revealed that it is mostly linked with increased replication efficiencies and severe SARS-CoV-2 infection as the patients containing this mutation had high viral load and two patients succumb to death.

## Discussion

A second wave of COVID-19 has grappled India, crossing 3.5 lakhs new cases per day with alarming death toll. An urgency persists to understand the clinical, epidemiological, immunological and genomic characteristics of the virus for correlating with the pathophysiology of SARS-CoV-2.

The current investigation delineates and compares the clinical manifestations, laboratory findings, immunological responses, genome sequences and treatment regimen of SARS-CoV-2 patients. Six asymptomatic and six symptomatic patients infected with this virus were studied for various parameters to understand the pathophysiology of this virus infection.

Analysis of the biochemical parameters highlighted the increased levels of CRP, LDH, serum SGPT, serum SGOT and ferritin in symptomatic patients indicating that patients with higher levels of few biochemical parameters might experience severe pathophysiological complications after SARS-CoV-2 infection. This was also observed that symptomatic patients were mostly with one or more comorbidities, especially diabetes (66.6%). Surprisingly the virological estimation revealed that there was no significant difference in viral load of OP samples between the two groups. This suggests that the viral load in OP sample does not correlate with disease severity and both asymptomatic and symptomatic patients are equally capable of transmitting the virus. Whereas, viral load was higher in plasma and serum samples of symptomatic patients suggesting that the development of clinical complications is mostly associated to high viral load in plasma and serum. This also demonstrated that the patients with high viral load in plasma and serum samples were found to develop sufficient amounts of antibodies (IgG, IgM and IgA). Interestingly, the levels of 7 cytokines (IL-6, IL-.1α, IP-10, IL-8, IL-10, IFN-α2, IL-15) were found to be highly elevated in symptomatic patients, while three cytokines (soluble CD40L, GRO and MDC) were remarkably higher in asymptomatic patients. Therefore, this data suggest that cytokines and chemokines may serve as “predictive indicator” of SARS-CoV-2 infection and contribute to understand the pathogenesis of COVID-19. The whole genome sequence analysis revealed that the current isolates were clustered with 19B, 20A and 20B clades, however acquired 11 additional changes in Orf1ab, spike, Orf3a, Orf8 and nucleocapsid proteins. The data also confirmed that the D614G mutation in spike protein is mostly linked with severe SARS-CoV-2 infection as two patients with this mutation passes away.

Patients with comorbidities displayed an increased severity in disease manifestation with some cases resulting in death. This observation was in accordance with previous report where mortality as well as severity of the illness were increased for the COVID-19 infected individuals with diabetes as compared to the non-diabetic patients (10). This study further confirms that the symptomatic patients had higher level of CRP and ferritin than the asymptomatic patients concurrent with previous reports (11,12). It was observed earlier that the viral load in nasal and throat swabs samples were nearly same in the asymptomatic and symptomatic patients. The current study also confirms the above observation suggesting that asymptomatic and symptomatic patients are equally contagious (13,14). In contrast, the viral loads in plasma and serum samples were associated with clinical complications that is consistent with other reports (15,16)

Additionally, the estimation of COVID-19 specific IgM, IgG and IgA antibodies in SARS-CoV-2 patients indicated that patients with high viral titer developed sufficient antibodies. A similar observation was reported in a study on patients of Hubei Province, China (15).

In this study, only one strain (with >10^7^ copies/mL) could be isolated successfully. This demonstrates that the viral load was insufficient to successfully isolate the virus from other samples. A previous study also reported the difficulty in isolation of the virus when viral load is <10^6^ copies/mL (17). This explains the inability in isolating the virus from the remaining samples.

Symptomatic patients showed higher levels of IP-10, IL-8, IL-10, IFN-α2, IL-15, IL-6 and IL-1α, whereas levels of soluble CD40L, GRO and MDC were higher in asymptomatic patients. In addition, IL-10 and IL-6 have been associated with severe disease manifestations. While IP-10 is considered to be a marker of ongoing inflammation, which in some cases predicts the mortality of the patients (18–21). Along with that, higher levels of IFN-α2, IL-15, IL-1α, IL-8 in the symptomatic patients indicate ongoing inflammation (22,23). The levels of CD40L, GRO and MDC were higher in the asymptomatic patients suggesting that they might have gone through immune suppression and repair phenomenon (24–26). Previous reports have shown significant difference in the expression levels of MCP-3, IL-1β, IL-17A, IL-12 p70, eotaxin and TNF, while in current study there was no significant difference observed as small sample size was a limitation(18).

Symptomatic patients showed higher levels of IgE, IgG2, IgG3 and IgG1 serotypes, as compared to asymptomatic patients. IgE has already been reported to be high in COVID-19 patients particularly those with Type II Diabetes. Out of 6 symptomatic patients in this study, 4 were suffering from type II Diabetes and had enhanced IgE levels (22,27). Overall increase in IgG1, IgG2 and IgG3 responses in symptomatic patients explains the increase in inflammatory immune responses(22). Since IgE was high in these patients there was no significant difference in IgG4 level as they compete for fixation sites in basophils and mast cells (28). There was no difference in IgM and IgA levels of the two groups suggesting that patients were at early stage of infection, as peak of virus specific IgM is developed approximately 14–28 days after the onset of symptoms (29).

The whole genome sequencing and phylogenetic analysis revealed that the current isolates were clustered in 19B, 20A and 20B clades, however there were several additional unique mutations in different genes as compared to the Wuhan strain. Surprisingly, most of the changes were observed in Orf1ab region. In India, these three clades were predominant in the months from May to July, 2020 (30). It was observed in this study that the three symptomatic patients with high viral load and among them two patients who had severe disease symptoms and died, were infected with the virus possessing the D614G mutation in the spike protein. This strengthens the previous hypothesis that the infectivity rate and enhanced transmission of SARS-CoV-2 is associated with the D614G mutation (9,31).

This study has indicated several interesting and significant outcomes, nonetheless with certain limitations. Primarily, all the clinical findings were correlated with viral RNA cycle threshold (Ct) value, which is capable of detecting dead virus particles also. Furthermore, the sample collection timing/interval was irregular and was solely dependent on the physician’s judgement and patient’s condition. Finally, given the small sample size, the results must be interpreted with caution and validated in a larger cohort to fortify these conclusions.

In summary, this investigation highlights the ability of both asymptomatic and symptomatic patients to transmit the virus equally. This also demonstrated that the D614G mutation is mostly associated with higher virus replication efficiency and severe SARS-CoV-2 infection. This data also suggest that the enhanced levels of inflammatory markers such as CRP and ferritin can be predictive biomarkers for critical condition of patients. Taken together, these results will contribute to a better understanding of the pathophysiology of SARS-CoV-2 infection and thereby advance in the implementation of effective disease control strategies.

## Data Availability

Whole genome sequencing data are fully available without restriction.

https://www.gisaid.org/

## ETHICS STATEMENT

The studies involving human participants were reviewed and approved by the Institutional Human Ethics Committee, Institute of Life Sciences. Written consent informed to participate in this study was provided by the participants’ legal guardian/next of kin.

## Author’s contribution

SC and SaC had designed the experiments. RS, SaC, SoS, ShS, SaS GB, and AR had collected the clinical specimens and demographic data. SaC, AD and SSK conducted the virological assays and RT-PCR. SoS and SaC had performed the bioplex and isotyping assays. SaC, AD and SoS had performed ELISA experiment. AJ and EL had prepared the samples for the whole genome sequencing. SC, SaC and AD analysed the virological data. SoS and SD analysed the immunological data. AG, SR and SW conducted the genomic data analysis. SC, SaC, AD, SoS and SSP had drafted the manuscript. AS, RD, SS, TKB, GHS, PP, SR, SD, RS, SC and AP reviewed and edited the manuscript.

## Author’s Agreement

This is to certify that all authors have seen and approved the final version of the manuscript being submitted. They warrant that the article is the authors’ original work, hasn’t received prior publication and isn’t under consideration for publication elsewhere.

## Data Availability

In the data availability statement the name of the Online repository is GISAID (https://www.gisaid.org/).

## Acknowledgement

We gratefully acknowledge ILS Biorepository for providing the SARS-CoV-2 samples collected from hospitals for this study. We will like to thank Dr. Subhasis Chattopadhyay, for his valuable suggestions in immunology section. We also acknowledge Mr. Paritosh Nath for collecting the samples from hospital, Mr. Jeky Chanwala for RNA isolation, Mr. Deepak Singh for data entry, Ms. Swati Madhulika and, Ms. Manasi Priyadarshini for helping in RT-PCR.

